# Understanding attrition: Feasibility and acceptability of a behavioral intervention for dementia care partners

**DOI:** 10.64898/2026.07.22.26357171

**Authors:** Allison Lindauer, Kristin G. Cloyes, Nathan Dieckmann, Christina Zonker, Heather Franklin, Susan Rosenkranz, Alex Speers, Michelle Kinsella, Christine McClure, Keely Young, Aimee Mooney

**Affiliations:** Department of Neurology, Oregon Alzheimer’s Disease Research Center, Oregon Health & Science University, Portland, Oregon, USA; School of Nursing, Oregon Health & Science University, Portland, Oregon, USA

## Abstract

**INTRODUCTION:** Behavioral intervention studies for family care partners for those with dementia need to be both feasible and acceptable in order implement and complete the investigative work. Our study, Tele-STELLA (Support via Technology: Living and Learning with Advancing dementia), was as single arm clinical trial completed in 2025. While 188 care partners enrolled, the attrition rate was high (44%). Here we describe the feasibility, acceptability, fidelity processes and preliminary efficacy of Tele-STELLA.

**METHODS:** Quantitative measures were used to assess care partner burden, study participation, feasibility and acceptability. Our weekly survey assessed the prevalence of adverse events. Qualitative methods paralleled our quantitative findings, in that care partners generally found the study acceptable, but dementia progression and life demands made participation difficult for some.

**RESULTS:** The total attrition rate was 44% but was attenuated by increasing the sample size. This adequately-powered study found that the intervention significantly reduced burden. Overall, care partners found the study feasible, but care demands made participation difficult for 28 of the care partners resulting in their withdrawal. In addition, 21 care recipients died, and thus their care partners had to be removed from the study. Protocol deviations and losing care partner contact also added to the attrition rate.

**DISCUSSION:** Our findings reveal that, even in the later stages of dementia, care partners are willing to participate in intervention research. However, care demands and death can affect the sample size. Our data and recommendations for future studies will inform caregiving scientists in designing behavioral interventions in late-stage dementia.

## 1 Background

Over 50% of care partners for those with dementia have at least one chronic medical condition and almost 60% of them report high emotional stress due to caregiving. And while mortality rates for other diseases such as heart disease and cancer are decreasing, deaths from dementia continue to rise.^1,2^ Together, these data suggest that participation in behavioral intervention research would be challenging for families living with dementia, especially for those in the advanced disease stages. Tele-STELLA (Support via Technology: Living and Learning with Advancing dementia) is an intervention designed to facilitate effective management of behavioral symptoms of dementia.^3^ Family care partners for those with moderate to severe stage dementia were recruited for this multi-component intervention. However, in line with the above statistics, the attrition rate was high (44%).

Attrition rates in behavioral research are affected by feasibility and acceptability. Feasibility is recognized as the capability of successfully implementing and completing a study, while acceptability focuses on study participants’ and staff members’ recognition of the study as suitable and satisfactory. If a study is feasible, resources (e.g., time, funds, etc.) are used appropriately to test important concepts and interventions. If a study is perceived as acceptable, participants are more likely to complete study components and study staff members are more likely to adhere to the protocol.^4,5^ Importantly, studies that are feasible and acceptable can mature into scalable interventions that address the needs of families caring for persons with dementia.^3,6^

Here we report results on preliminary efficacy, feasibility, acceptability, and the fidelity assessment of Tele-STELLA. We discuss study challenges and the adaptations we implemented to maintain study integrity and meaningful results.

## 2 Methods

This prospective, single-arm clinical trial tested Tele-STELLA, a behavioral intervention for adults caring for a family member with dementia. The objectives of this paper are to report on the primary aim of this NIH Stage 1 intervention,^6^ that is, to test feasibility and acceptability of Tele-STELLA, as well as the second aim, which addressed preliminary efficacy by assessing the impact of the intervention on care partner burden.^3^

The Tele-STELLA telehealth-based intervention teaches care partners how to address the behavioral symptoms associated with advancing dementia using the ABC (Activator, Behavior, Consequences) approach (see Lindauer et al., 2021^3^). Care partners participated in the study, from their homes, via HIPAA-compliant videoconferencing. We tested the intervention in two components: Nova and Constellation. In Nova, the main component, caregivers are taught by the interventionist (“Guides”) to identify the most bothersome care recipient behaviors; they are then instructed in a methodological approach to address these behaviors. Nova took place over 8 eight weeks, with four weeks of 1-hour individual instruction and four of 1- hour small group sessions.

Constellation, the second component of Tele-STELLA, is an 8-week booster dose of the same curriculum delivered during Nova, but after Nova and in a larger group format with up to 12 care partners and two Guides. While Constellation was a required component of Tele-STELLA, care partners had up to one year to join a Constellation group after completing Nova. Constellation was not tested in our pilot work, thus testing this booster dose was an important exploratory component of the study.

Active participants included family members (“care partners”) who cared for a person with dementia (“care recipients”). While care recipients did not participate in any of the active components of Tele-STELLA, we collected qualitative and quantitative data about them. Further, we recognized our ethical responsibility of informing them of their privacy rights and confidentiality concerns, such as entering the background of their care partner’s video during an intervention session. Thus, care recipients were also consented and enrolled.

We aimed to recruit 150 care partners based on the original power calculation which found that 124 care partners were needed to provide 90% power to detect a 2-point reduction on the primary burden outcome measure, reactivity, on the Revised Memory and Behavior Problem Checklist (RMBPC).^7^ The pilot work testing the intervention had a 15% attrition rate, thus, our original goal was to recruit 150 care partners (and their 150 care recipients) into Tele-STELLA. We examined the success of our recruitment approach by tracking enrollments and screen failures on a weekly and quarterly basis, comparing progress to goals set at study start. Retention strategies are described elsewhere.^3^

To be included in the study care partners had to report at least two care recipient behavioral symptoms that upset them. Because behavioral symptoms of dementia are most common in the moderate to late stages of dementia, care recipients with mild dementia, described by the Alzheimer’s Association, were excluded.^8^ The care partner and care recipient did not need to live together, but both had to be alive during the study period. Care recipients receiving hospice care were included in the study.

Tele-STELLA is a technology-based intervention. All assessments and interventions in this study were completed by phone, videoconferencing, or electronically administered assessments. To facilitate ease of access and standardization, care partners who did not have a functional computer were loaned a laptop programmed for study-use only. Technology support was available by an experienced team member to any care partners that needed it, further limiting technology barriers to study participation for older adult and rural care partners.

### 2.1 Funding

Tele-STELLA was funded by the National Institute on Aging (NIA), R01AG067546, with other support from the Oregon Alzheimer’s Disease Research Center, NIA P30AG066518.

### 2.2 Consent Statement

The study was approved initially by Western IRB, a single IRB (#1303009). In 2022, IRB oversight was transferred to Oregon Health & Science University (IRB#22288). Tele-STELLA is registered with Clinicatrials.gov (#NCT04627662; 11/08/2020). All participants (care partners and care recipients) were consented and enrolled in Tele-STELLA. If the person with dementia was not able to consent, the legally authorized representative consented for them. If the care recipient did not consent to study participation, the care partner could not join the study.

### 2.3 Safety

This was a minimal risk study. However, because the care recipients were in the later stages of dementia, we anticipated that the number of urgent care visits, hospitalizations, deaths, and other adverse events would be high. Working the NIA program officers, we developed a Data Safety Monitoring Plan that recognized these events as “expected.” All adverse events, for both care partners and care recipients, were reported quarterly to the Tele-STELLA Safety Officer, the OHSU IRB, and the NIA. Severe adverse events were reported to these entities immediately.

### 2.4 Measures and Analyses

#### 2.4.1 Efficacy

Mixed-effects modeling was used to assess preliminary efficacy of Tele-STELLA. The primary burden outcome measure was care partner reactivity to standardized behavioral symptoms of dementia, as measured on the RMBPC.^7^ The RMBPC was administered five times via electronic surveys ^9^ over the course of participation. Care partners also completed a weekly “Orbit” survey that collected weekly data on adverse events, including urgent care and hospital use.

#### 2.4.2 Feasibility

The Formal Feasibility Study framework informed the Tele-STELLA feasibility assessment.^5^ This included examination of recruitment efficacy as well as attrition and completion rates of both Tele-STELLA intervention components (Nova and Constellation) and surveys. The completion rates of Tele-STELLA study components and reasons for not completing were assessed. Administrative lag from consent to Nova initiation was also examined to evaluate whether delays in intervention initiation were associated with non-completion.

We defined attrition as any loss of participants from Tele-STELLA.^10^ Attrition was considered in relation to feasibility, acceptability, and fidelity to study protocol and divided into three categories (Table 1). Rates and percentages were tabulated. Predictions of attrition were analyzed using logistic regression models which included age, caregiver burden, study satisfaction, dementia severity, and adverse events (e.g., falls, illness, hospitalization). Separate and combined models examined caregiver and care recipient adverse events occurring during the study.

**Table 1.** Tele-STELLA Attrition (Non-Completer) Definitions.

| **Participant Lost to Follow-Up** | **Participant-Initiated Withdraw** | **Investigator-Initiated Removal** |
| --- | --- | --- |
| Participant could not be reached after multiple contact attempts and the reason for stopping was unknown | The care partner chose to stop participation and/or clearly communicated a personal reason for stopping the study | Investigator ends participation for protocol deviations, safety, eligibility, administrative, or study-related reasons |

Care partners were queried about feasibility using the Tele-STELLA Experience Survey. Care partners responded to items on a 5-point scale, with 1= “strongly disagree” to 5 “strongly agree” to statements such as, “It was easy for me to attend the Tele-STELLA visits.”

#### 2.4.3 Acceptability

As with the feasibility assessment, acceptability was examined using quantitative and qualitive strategies. Guided by the Theoretical Framework of Acceptability, acceptability was assessed retrospectively by examining care partner attitudes, burden of participation, and perceived effectiveness using quantitative and qualitive strategies.^4^ Care partners were queried about acceptability using the Tele-STELLA Experience Survey. Care partners responded to items on a 5-point scale, with 1= “strongly disagree” to 5 “strongly agree” to statements such as, “Overall, I was satisfied with the Tele-STELLA experience.”

#### 2.4.4 Fidelity

A fidelity plan for study staff was implemented at study start, modeled on Teri et al.’s approach.^11,12^ As the study progressed, the fidelity plan was revised to improved processes and data collection. Procedural, theoretical, documentation, and treatment fidelity outcomes were assessed. Procedural fidelity reflected completion of required intervention components. Theoretical fidelity reflected interventionist adherence to the intervention framework and use of core intervention strategies. Documentation fidelity reflected completeness and timeliness of session documentation, and treatment fidelity assessed the update of the lessons by the participants.^13^

Procedural, theoretical, and documentation fidelity was assessed using the Protocol Adherence Checklist (PAC), which included items completed by Guides and items completed by the Fidelity Assessment Team (FAT). The FAT was composed of study investigators and subject matter experts. Video recordings and transcripts of intervention sessions were reviewed by the FAT to assess required elements of the protocol and alignment with the intervention framework. Associated documentation was also reviewed to assess completeness and timeliness of required data capture. Treatment fidelity was assessed using the Tele-STELLA Experience Survey, and qualitatively via focus groups.^13,14^

After the revised fidelity procedures were refined, a random sample of 30% of sessions from each cohort was selected for fidelity review. Inter-rater reliability was evaluated for Nova mid-study cohorts using Cohen’s kappa. Protocol adherence reliability was calculated across items completed by both the FAT and Guides, while theoretical adherence reliability was calculated using assessor-only items from the PAC. Documentation adherence was assessed using the proportion of checklist items coded as missing. Timeliness of documentation was assessed using the average number of days between the intervention session and completion of the PAC. Fidelity data were summarized across cohorts to examine changes following iterative refinement and implementation of the revised fidelity procedures.^15^

#### 2.4.5 Focus Groups

Along with quantitative assessments, feasibility, acceptability, and fidelity were assessed qualitatively using focus groups, convened via videoconferencing. A directed content analysis approach was used to summarize and describe the data.^16^ Synthesis of feasibility findings was informed by the Formal Feasibility Study framework,^5^ while synthesis of acceptability findings was guided by and the Theoretical Framework of Acceptability.^4^

Overall, qualitative and quantitative data were analyzed using a parallel analytic approach, meaning data collection methods were independent of each other, but results mirror the two approaches and informed the overall findings.^17^

## 3 Results

### 3.1 Participants

Recruitment for Tele-STELLA started in May 2021 and the last participants completed the intervention in July 2025. Participants were initially recruited from four NIA Alzheimer’s Disease Research Centers. Despite contractual agreements and dedicated team effort, recruitment of disproportionately represented care partners fell short of expectations. In 2023, with NIA approval, recruitment efforts widened to include care partners from across the US. The total number screened for Tele-STELLA was 406. There were 233 screen failures, of these,158 (68%) met exclusion criteria, with these top three reasons: care partners being too busy (n=45, 28%), care recipients having mild dementia (n=35, 22%), or reporting a lack of upsetting behaviors (n=29, 18%) (Figure 1).

**Figure 1.**
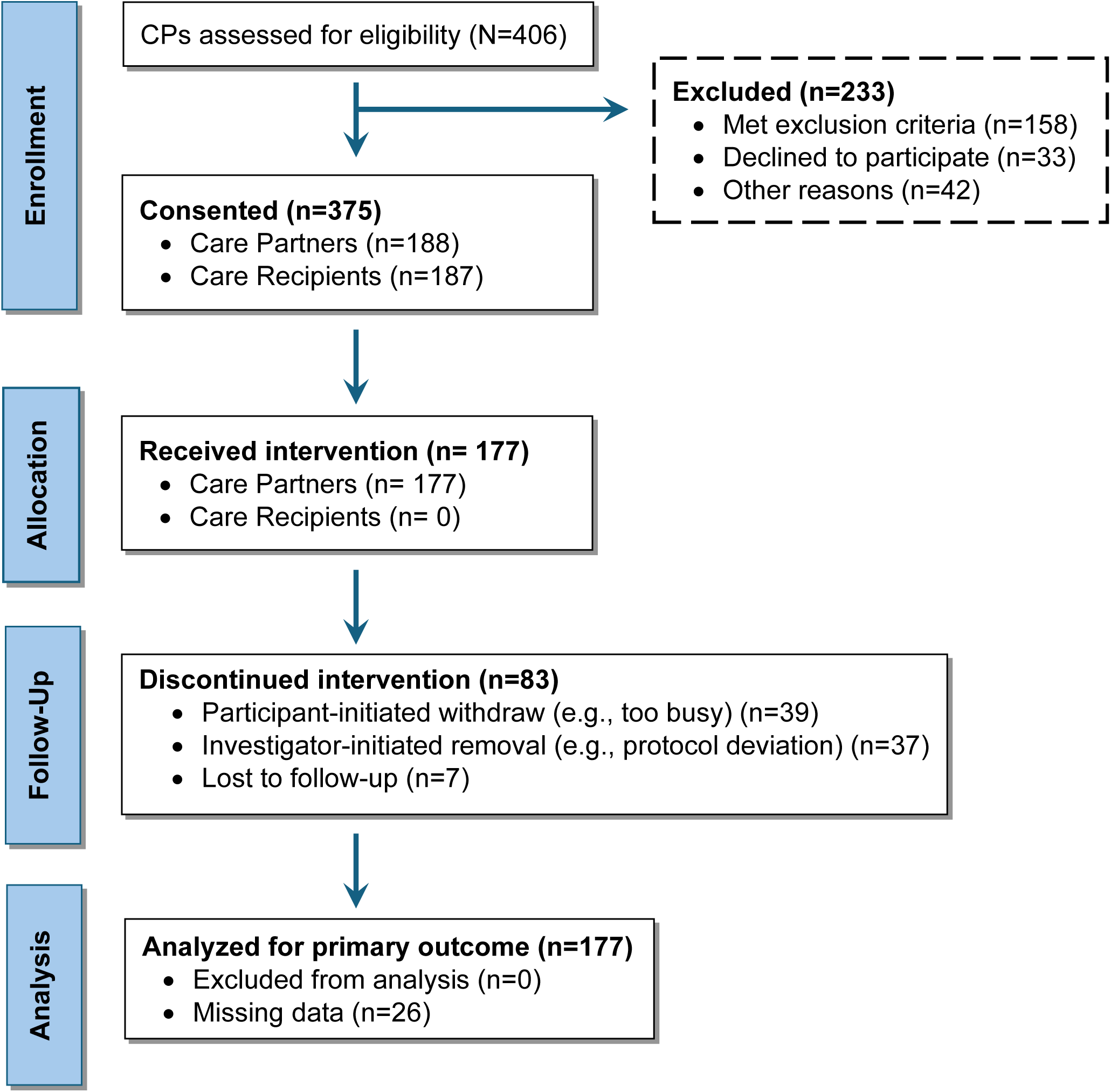
CONSORT Flow Diagram: Tele-STELLA Participant Engagement

Approximately half of the required sample of 124 care partners (determined by the power analysis) was enrolled by 2022. However, our attrition rate was about 30%. Working with the NIA program officer, we increased the projected sample size to 200 mid-study. The final sample included 188 care partners and 187 care recipients (one had two care partners), with 83 non-completers (Tables 2 and 3). Those that completed the study did not differ significantly that did not with regard to age, race, gender, education, dementia stage or baseline RMBPC reactivity score. Of the 161 care partners that completed Nova, 17 did not complete post-assessments, leaving 144 care partners completing pre- and post-Nova assessments. With 144 completing post-Nova assessments, the study was adequately powered to identify significant change post-Nova.

**Table 2.** Completers vs. Non-Completers Demographics: Nova.

|  | **Nova completer**<br>**(N=161)** | **Nova non-completer**<br>**(N=27)** | **P-value** |
| --- | --- | --- | --- |
| **Age** |  |  |  |
| Mean (SD) | 65.2 (11.3) | 61.6 (15.8) | 0.389 |
| Median [Min, Max] | 66.1 [30.7, 87.8] | 62.1 [38.2, 81.9] |  |
| Missing | 0 (0%) | 11 (40.7%) |  |
| **Race** |  |  |  |
| Non-White | 21 (13.0%) | 6 (22.2%) | 0.336 |
| White | 140 (87.0%) | 21 (77.8%) |  |
| **Gender** |  |  |  |
| Male | 43 (26.7%) | 5 (18.5%) | 0.559 |
| Female | 117 (72.7%) | 21 (77.8%) |  |
| Missing | 1 (0.6%) | 1 (3.7%) |  |
| **Education** |  |  |  |
| Mean (SD) | 16.3 (2.13) | 16.0 (2.43) | 0.488 |
| Median [Min, Max] | 16.0 [12.0, 20.0] | 16.0 [12.0, 20.0] |  |
| **Dementia Stage** |  |  |  |
| Moderate | 142 (88.2%) | 22 (81.5%) | 0.512 |
| Severe | 19 (11.8%) | 5 (18.5%) |  |
| **RMBPC pre-Nova** |  |  |  |
| Mean (SD) | 40.6 (15.5) | 42.6 (14.2) | 0.604 |
| Median [Min, Max] | 40.9 [0, 81.6] | 41.6 [20.0, 66.7] |  |
| Missing | 1 (0.6%) | 11 (40.7%) |  |

**Table 3.** Completers vs. Non-Completers Demographics: Constellation.

|  | **Constellation**<br>**completer**<br>**(N=112)** | **Constellation non-**<br>**completer**<br>**(N=16)** | **P-value** |
| --- | --- | --- | --- |
| **Age** |  |  |  |
| Mean (SD) | 65.1 (10.8) | 68.9 (10.0) | 0.176 |
| Median [Min, Max] | 66.4 [36.9, 86.4] | 73.3 [51.7, 83.1] |  |
| **Race** |  |  |  |
| Non-White | 16 (14.3%) | 2 (12.5%) | 1.000 |
| White | 96 (85.7%) | 14 (87.5%) |  |
| **Gender** |  |  |  |
| Male | 29 (25.9%) | 3 (18.8%) | 0.743 |
| Female | 82 (73.2%) | 13 (81.3%) |  |
| Missing | 1 (0.9%) | 0 (0%) |  |
| **Education** |  |  |  |
| Mean (SD) | 16.2 (2.15) | 16.8 (1.91) | 0.248 |
| Median [Min, Max] | 16.0 [12.0, 20.0] | 16.0 [12.0, 20.0] |  |
| **Dementia Stage** |  |  |  |
| Moderate | 105 (93.8%) | 13 (81.3%) | 0.213 |
| Severe | 7 (6.3%) | 3 (18.8%) |  |
| **RMBPC pre-Constellation** |  |  |  |
| Mean (SD) | 35.1 (16.2) | 34.2 (19.6) | 0.869 |
| Median [Min, Max] | 34.6 [1.41, 76.8] | 33.8 [6.26, 85.1] |  |
| Missing | 12 (10.7%) | 2 (12.5%) |  |

### 3.2 Efficacy

Among participants who completed the Nova intervention (N=144), mixed-effects models demonstrated significant reductions in RMBPC total reactivity from baseline to post-Nova (β = –4.47, p < .001) that were maintained at follow-up (one month post Nova) (β = –4.51, p < .001). Effect sizes were small-to-moderate (d ≈ –0.28).

Analysis of the full sample, including those who did not complete Nova, (n=177) found a significant reduction in total reactivity on the RMBPC ^7^ (β = –4.76, p < .001) and remained improved at follow-up (β = –4.63, p < .001), representing the largest overall effect (f² = 0.27, d = –0.29 to –0.30).

Mean total reactivity was slightly lower over Constellation timepoints (pre-Constellation mean 35.0, post-Constellation mean 33.2, follow-up mean 33.0), with continued between-person heterogeneity.

From a qualitative perspective, result findings from the focus groups suggest that care partners found the intervention helpful. Perceived usefulness was one of the strongest and most consistent signals. Participants described gaining practical tools for managing behaviors, reframing caregiving situations, and developing new ways to respond to challenges. One participant described the program as providing a “pathway” to think through situations and regain control during an “emotional tailspin”, while others emphasized that focusing on specific behaviors helped them “navigate that forest” of caregiving demands. Usefulness appeared to operate through both practical behavior management and cognitive reframing.

Overall, the quantitative findings signal that Tele-STELLA is effective in reducing care partner burden. These findings are complemented by the qualitative results suggesting the intervention was generally perceived as useful.

### 3.3 Feasibility

Of the 188 total care partners enrolled in Tele-STELLA, n=161 (86%) completed Nova, 113 (60%) completed Constellation, and 105 (56%) completed the full study (Tables 2, 3). Over the course of the study (from 2021-2025) completion of the weekly survey averaged 74%, ranging from the lowest in 2023 at 67% to the highest in 2021 at 87%. On average, 93% of the six longer assessments were completed. The lowest completion rate was for the 1-month post Nova intervention, at 85%.

Of the care partners that enrolled in Tele-STELLA, n=83 (44%) did not complete the study due to a) care partner-initiated withdraws (n=40, 48% of all non-completers), b) investigator-initiated removals (n=36, 43%), or lost to follow-up (n=7, 8%) (Figure 2.)

**Figure 2.**
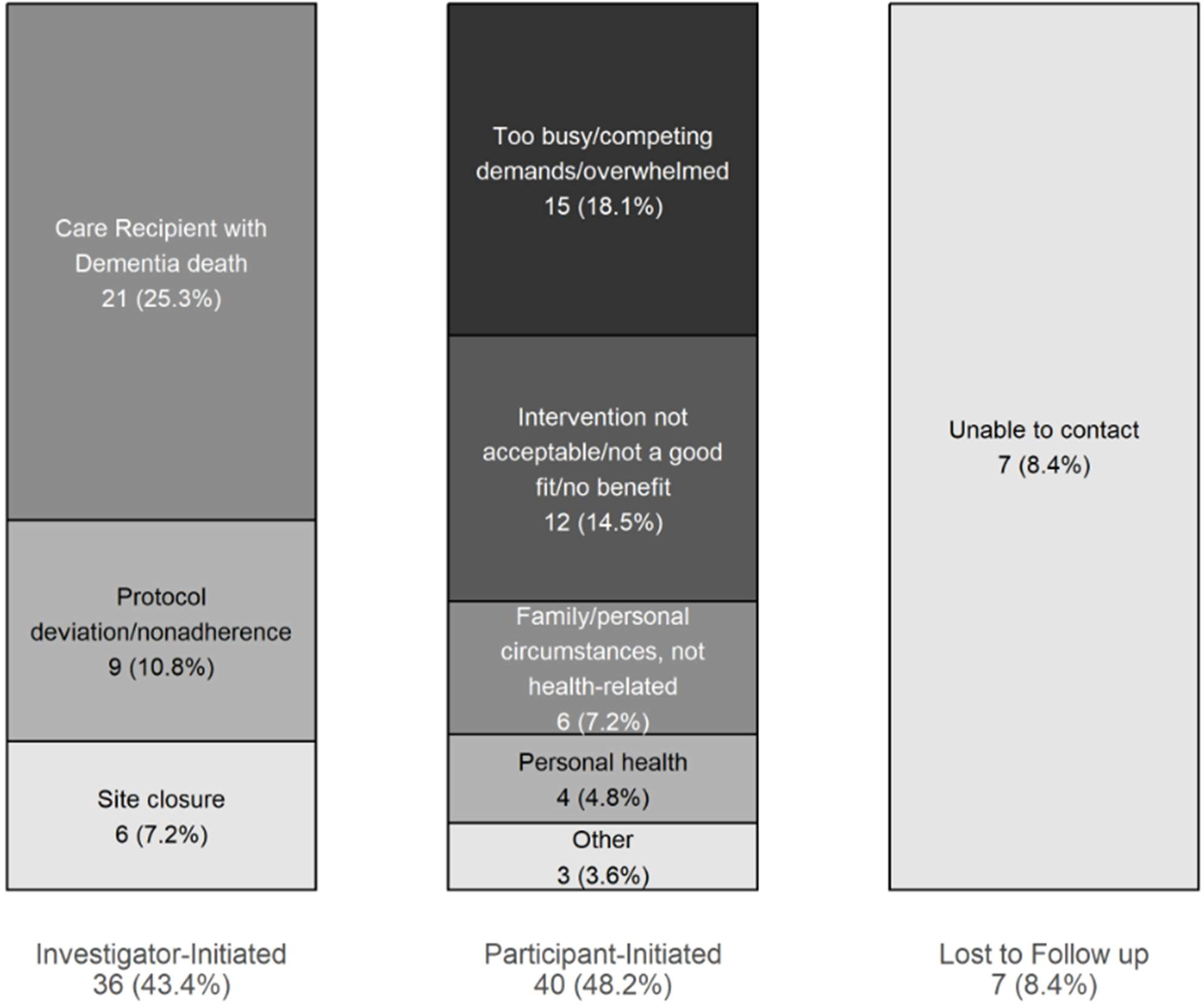
Attrition Reasons by Non-Completion Group (N=83)

Care partner-initiated withdraws were due to factors associated with increasing care burden, e.g. being too busy, competing demands, and/or health issues (n=28; 34% of attrition). A smaller group reported that the study was not a good fit or did not benefit them (n=12; 14%).

Investigator-initiated removal occurred primarily due to care recipient death (n= 21; 25%). Of the 21 deaths, the majority (67% of all deaths) occurred after Nova, but before the start of Constellation; one death occurred prior to the care partner starting the intervention (Nova) (Figure 3). The majority of the deaths (n=16, 76% of deaths) occurred in care recipients in the moderate stages, with 5 deaths occurring in those in the advanced stages. Care recipients who died during the study were significantly older than those who did not die. Among participants who withdrew, the mean age of care recipients was 85.2 years (SD = 7.0) for those who died, compared with 79.5 years (SD = 8.4) for those who did not die (p = .004). A similar difference was observed in the full sample (85.2 vs. 80.2 years, p = .006).

**Figure 3.**
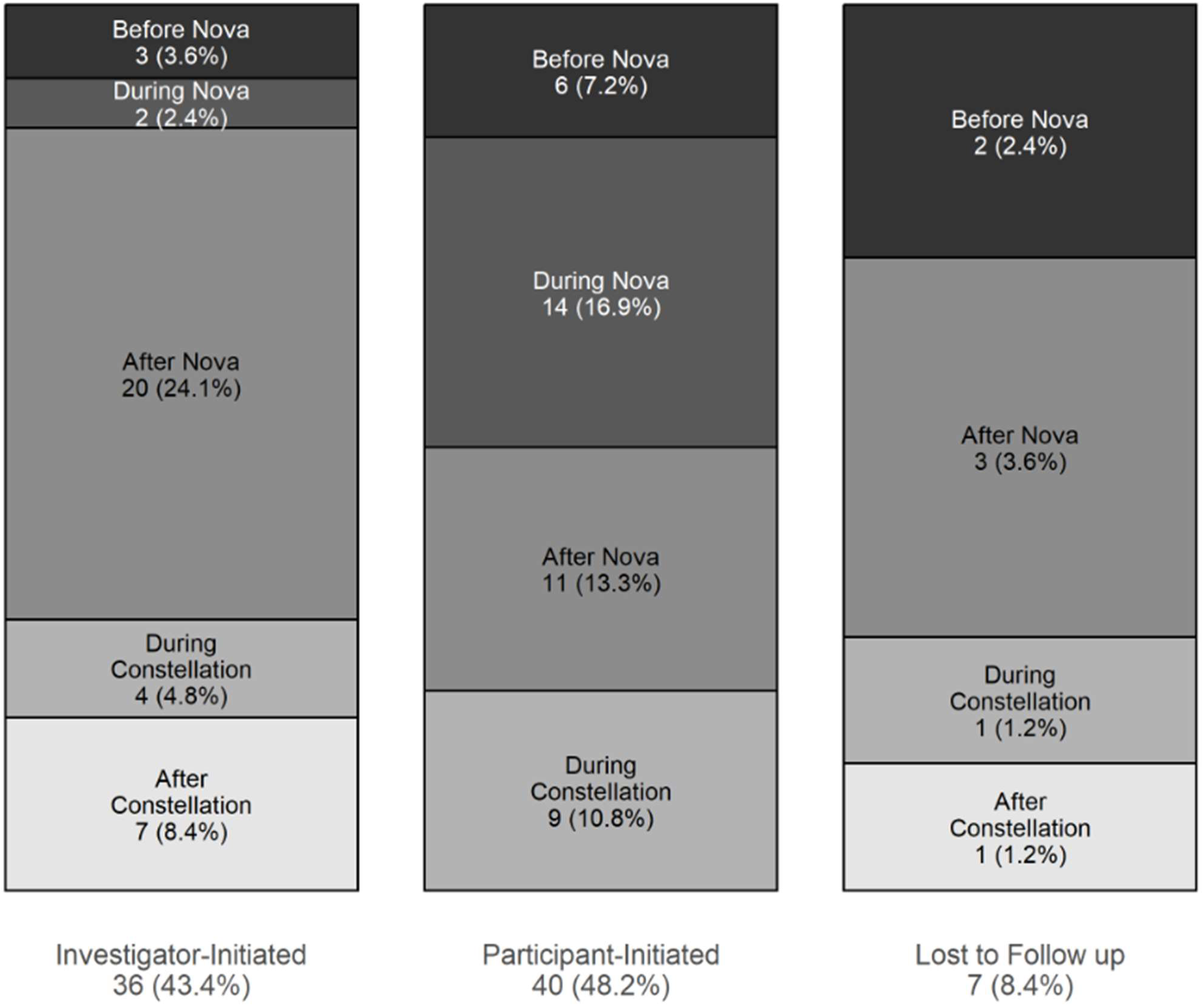
Attrition Timing by Non-Completion Group (N=83)

Predictors of withdraw revealed that, overall, age, care partner burden and study satisfaction, did not predict withdrawal. Severe dementia increased odds of withdrawing nearly threefold (OR=2.92, p=0.09).

The weekly Orbit survey identified 198 adverse events, and the number of adverse events in the care recipient predicted withdrawal, with each event increasing the odds by 29% (OR=1.29, *p*=0.07). Among participants completing Nova, greater care recipient non-death adverse events during Nova were associated with earlier non-completion. In adjusted Cox proportional hazards models, each additional care recipient non-death adverse event during Nova was associated with 69% higher hazards of non-completion (adjusted HR = 1.69, 95% CI: 1.09–2.62).

Severe dementia independently predicted earlier withdrawal (adjusted HR = 2.57, 95% CI: 1.35–4.92). Kaplan–Meier analyses demonstrated a dose-response pattern, with participants experiencing two or more care recipient non-death adverse events during Nova demonstrating the earliest withdraw following Nova completion. Caregiver non-death adverse events were not independently associated with withdraw or time-to-withdraw.

Technological challenges were few and were not associated with withdrawal. The Tele-STELLA experience survey found that, given the option in future study of in-person or via telehealth, 88% of the participant preferred the telehealth format. Administrative lag from consent to Nova initiation was not associated with overall non-completion, although participants who withdrew prior to starting Nova had substantially longer delays before intervention initiation.

Examination of the study timeline revealed that most care partner-initiated stoppage occurred during Nova, due to either participant-initiated withdrawal or lost to follow-up. Most investigator-initiated removals occurred after Nova, most often due to the death of the care recipient (Figure 3).

Feasibility was also examined with the Tele-STELLA Experience Survey. Of the 143 care partners who completed Nova, 142 responded to the experience survey and 97 completed it after Constellation. The mean rating was above 4 (“agree”) on all feasibility items (Table 4).

**Table 4.** The Tele-STELLA Experience Survey: Feasibility.

|  | **Post-Nova**<br>**(N=142)** | **Post-**<br>**Constellation**<br>**(N=97)** |
| --- | --- | --- |
| **It was easy for me to attend the Tele-STELLA visits.** |  |  |
| Mean (SD) | 4.14 (0.819) | 4.45 (0.732) |
| Median [Min, Max] | 4.00 [1.00, 5.00] | 5.00 [2.00, 5.00] |
| **It was easy for me to find a safe activity for my family member with dementia during the training sessions.** |  |  |
| Mean (SD) | 4.06 (0.936) | 4.01 (1.060) |
| Median [Min, Max] | 5.00 [2.00, 5.00] | 4.00 [1.00, 5.00] |
| **I had good technical support from the study team.** |  |  |
| Mean (SD) | 4.54 (0.646) | 4.65 (0.498) |
| Median [Min, Max] | 5.00 [1.00, 5.00] | 5.00 [3.00, 5.00] |
| **I could easily see and hear the Tele-STELLA Guide(s) and other participants.** |  |  |
| Mean (SD) | 4.30 (0.833) | 4.34 (0.836) |
| Median [Min, Max] | 4.00 [1.00, 5.00] | 5.00 [1.00, 5.00] |
| **It was easy to connect with Tele-STELLA using my computer or other device.** |  |  |
| Mean (SD) | 4.46 (0.756) | 4.52 (0.690) |
| Median [Min, Max] | 5.00 [1.00, 5.00] | 5.00 [2.00, 5.00] |

Focus group findings informed the quantitative results. Four groups were made up of a subsample of Tele-STELLA participants who completed the study (n=25). These four groups had a mean age of 61.7 years (SD = 8.8; range 36–80) and were predominantly female (80.8%). Care relationships were split between spouses/partners (53.8%) and adult children (46.2%). Care partners took part in the focus groups, on average, 30 months after consenting to Tele-STELLA (range 16-51 months). We conducted one focus group with care partners who withdrew prior to study completion (n=4), (n=4), mean age: 67.3 years (SD: 10.5, Range: 54-79). All were female, three were spouses, one was an adult child. These care partners attended the focus group, on average, 22 months after their participation ended (range 18-24 months).

NIA P30AG066518The feasibility findings from the focus groups aligned with five key themes: Time and Scheduling, Caregiving Demands/Caregiver responsibilities, Technology Access and Ease, Fit with Daily Life, and Participation Supports/Facilitators.

#### Time and Scheduling

Participants across groups described time as a central feasibility constraint, but not as a simple scheduling issue. Rather, time was experienced as already fully occupied by caregiving responsibilities, requiring participants to actively “make time” for sessions. This often involved prioritization, trade-offs, and, in some cases, feelings of guilt about stepping away from caregiving duties. Time constraints were therefore embedded within the broader structure of caregiving rather than functioning as an isolated barrier.

#### Caregiving Demands / Competing Responsibilities

Caregiving demands emerged as one of the most consistent feasibility challenges. Participants described dementia caregiving as unpredictable and continuous, with needs that could change rapidly and without warning. This made it difficult to commit to scheduled sessions or remain fully engaged once a session began. Several participants noted the need to keep the person with dementia occupied and calm while the care partner was in the Tele-STELLA sessions underscored the tension between participating and remaining responsible for care.

#### Technology Access and Ease

Technology-related challenges were present but did not appear to be a primary barrier to participation. When mentioned, issues such as connectivity or platform use were generally manageable and secondary to caregiving demands. Participants often framed the telehealth format as facilitating participation relative to in-person options, even if it did not eliminate other constraints.

#### Fit with Daily Life

Fit with daily life varied across participants and appeared to depend on the stage of dementia, length of time stage of dementia, length of time caregiving, relationship to the care recipient, and overall context. Some participants described the intervention as aligning well with their needs at a critical time, while others noted mismatches between the program structure and their caregiving situation. Differences in experience level or caregiving role (e.g., spouse vs. adult child) also influenced how well the intervention fit participants’ daily realities.

#### Participation Supports / Facilitators

Participants identified several factors that facilitated participation, including respite care, flexible scheduling, supportive study staff, and remote access. These facilitators often functioned as enabling conditions rather than conveniences, allowing participants to create protected time for engagement. The availability of support, whether formal or informal, appeared to be a key determinant of whether participation was feasible.

In summary, the Nova component was feasible for most care partners, but the booster Constellation was not, as evidenced by the high attrition rate post Nova. Care recipient death, competing demands, and schedule challenges contributed to the high non-completion rates. Based on these findings, future estimations of sample size will take these factors into account. We will prepare future protocols to be adaptive to attrition rates.^5^ Due high withdrawal rate, we will consider eliminating the booster Constellation component in future studies.

### 3.4 Acceptability

Tele-STELLA Experience Survey revealed that most of the care partners agreed or strongly agreed with the statement, “Overall, I was satisfied with the Tele-STELLA experience” (Table 5). Qualitative data was gathered from the focus groups (the same groups described above), findings fell into four categories: Satisfaction/Liking, Relevance / Appropriateness / Fit, Effort, Would Recommend / Continue.

**Table 5.**
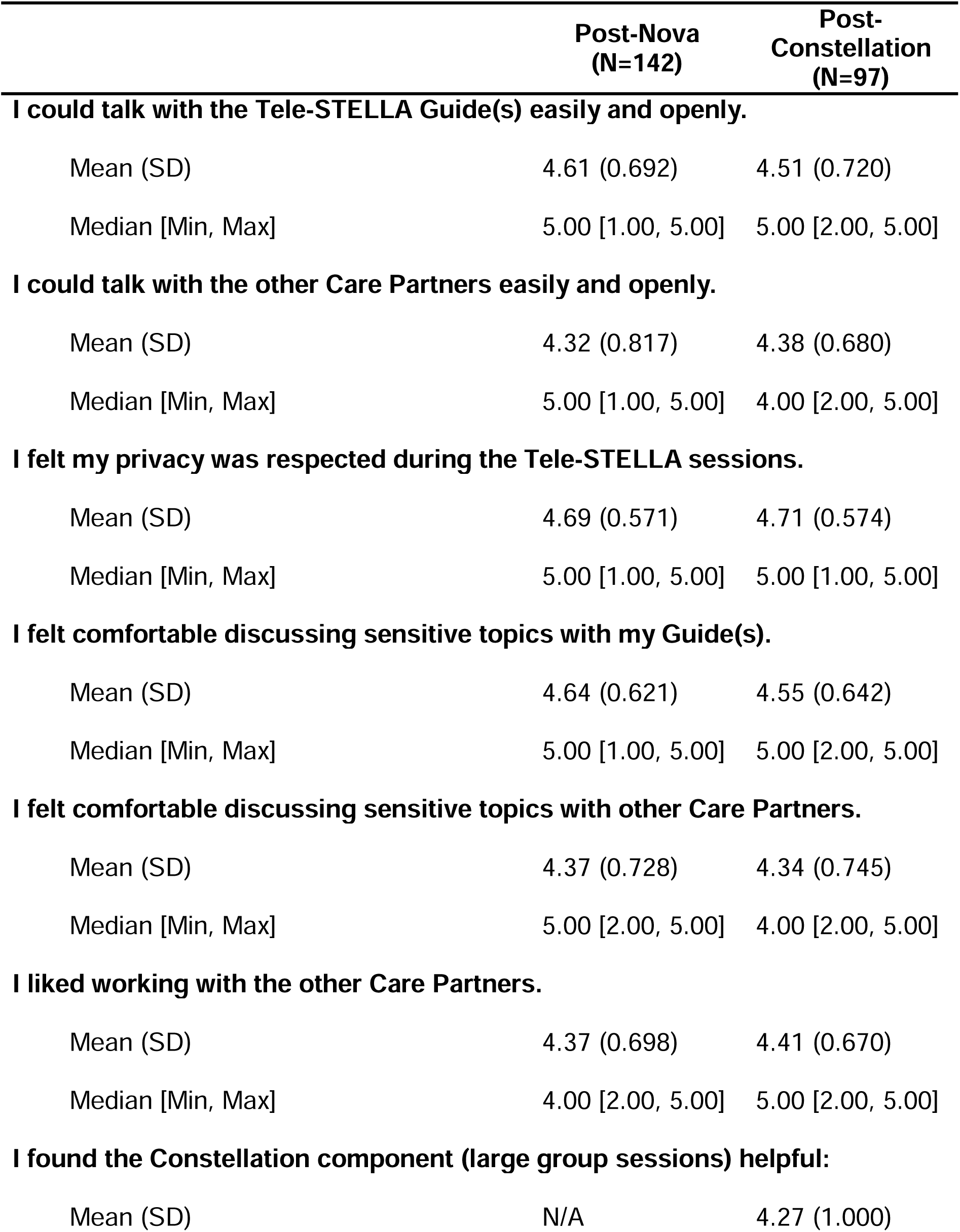
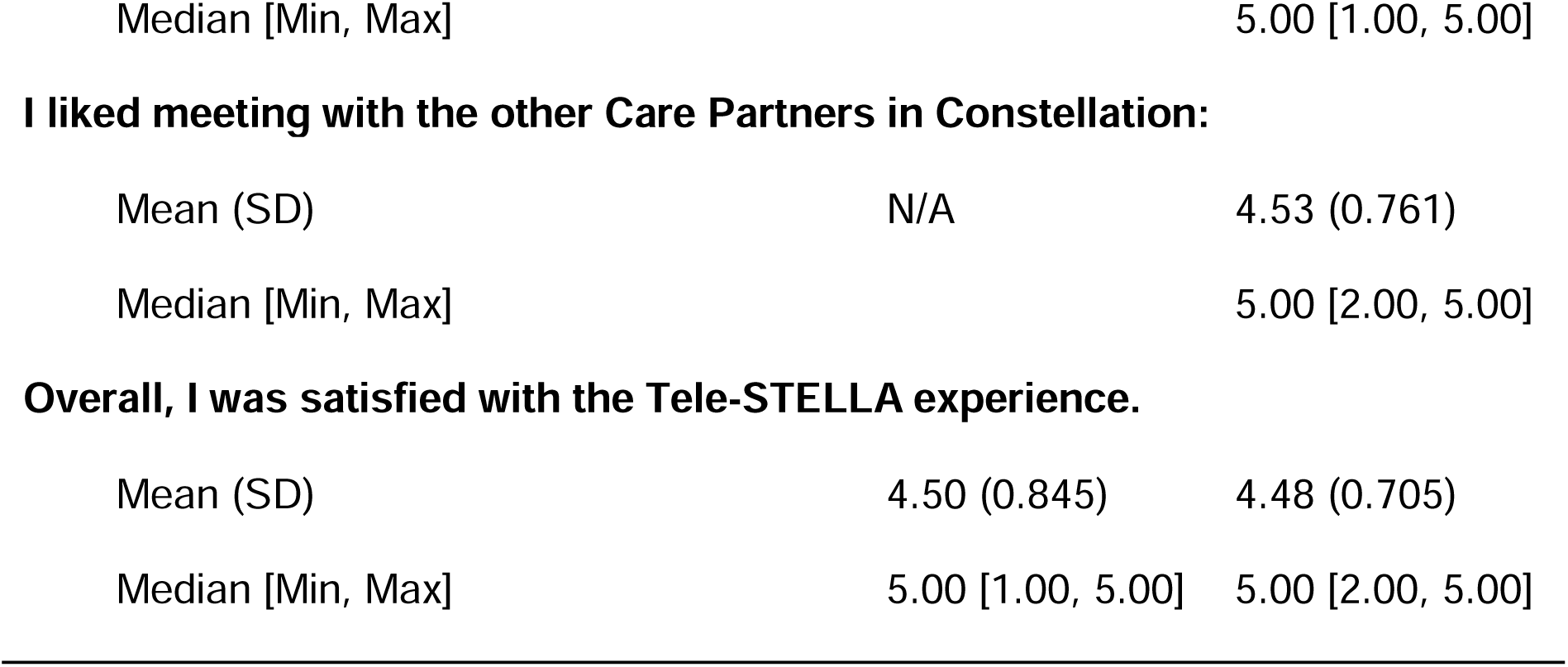
Tele-STELLA Experience Survey: Acceptability.

#### Satisfaction / Liking

Participants generally expressed positive overall reactions to Tele-STELLA, describing it as helpful, valuable, and meaningful. Satisfaction was often tied to specific aspects of the experience, including feeling supported, gaining insight, and having a structured space to discuss caregiving challenges. Participants emphasized the meaningful impact of the program, describing it as a “a rescue line” and “a life changer,” and noting that both strategies and connections with others “basically set [them] on a course that kept [them] upright.”

#### Relevance / Appropriateness / Fit

Relevance varied across participants but was strongest among those who were actively seeking support, education, or practical strategies. Many described the intervention as meeting an immediate need during a period of stress or uncertainty. At the same time, some participants noted that differences in caregiving context or group composition could limit applicability. Relevance appeared closely tied to caregiving stage and context, for example, one care partner described a mismatch because they had many years of experience in caregiving (“we were 15 years in… it just didn’t quite fit me”). Differences in caregiving roles also shaped fit. These patterns suggest that relevance is not uniform, but varies based on caregiving stage, relationship to the care recipient, and alignment between participants’ immediate needs and the structure of the intervention.

#### Effort

Participants identified elements of effort associated with written materials, structured exercises, and tasks that resembled “homework.” These components were sometimes experienced as effortful, particularly when participants were already managing emotional overwhelm and complex caregiving logistics. However, this effort did not appear to undermine overall acceptability, because participants often balanced these challenges with perceived benefits from other aspects of the intervention.

#### Would Recommend / Continue

Willingness to continue or recommend the intervention was reflected in participants’ behaviors and ongoing engagement. Participants described continuing to use intervention principles and maintaining connections with other group members, including ongoing “weekly calls together.” These patterns suggest sustained value beyond the formal intervention period.

In brief, these acceptability findings indicate that care partners appraised Tele-STELLA in a positive light. However, the intervention appears to be a better fit for care partners in the earlier years of caregiving. Future effort should consider the value of incorporating written exercises as part of the intervention.

### 3.5 Fidelity

Fidelity to the protocol was assessed across the entire study. Review of early cohorts identified variability in intervention delivery, alignment with the theoretical framework, and documentation practices. The initial fidelity approach identified inconsistencies in Guide adherence to the protocol as well as limitations in the fidelity assessment process, including low inter-rater reliability among assessors and missing data.^15^ After review of the first three cohorts, fidelity procedures were iteratively refined and phased in over subsequent cohorts.

The full fidelity monitoring approach was implemented and fully integrated mid-study. Revisions included modification of the Guide Manual, Guide re-orientation, refinement of the Protocol Adherence Checklist, assessor re-training, and creation of REDCap forms that consolidated protocol tasks, recommended Guide facilitation behaviors, and documentation requirements into one system.^9^ Definitions of essential Guide facilitation behaviors were clarified and incorporated into standardized checklists using binary present/absent ratings to improve consistency in assessment of theoretical fidelity. These changes simplified protocol documentation, improved efficiency for both Guides and fidelity assessors, and supported more consistent monitoring. To maintain protocol adherence and reduce drift over time, the team also implemented a structured agenda for weekly Guide meetings, email reminders highlighting a “Protocol Tip of the Week,” and quarterly fidelity updates for the full study team. Together, these updates strengthened the fidelity infrastructure and supported more consistent intervention delivery across later cohorts.

Fidelity outcomes generally improved across cohorts, with improvements becoming more apparent following the complete implementation of fidelity enhancements mid-study. Protocol adherence reliability increased from a mean kappa of 0.76 mid-study to 0.84 in the last sessions. Values remained in the substantial-to-almost-perfect agreement range from mid-study to completion. Theoretical adherence reliability showed a more pronounced improvement, increasing from 0.69 mid-study to 1.00 in later sessions. Documentation adherence was consistently high across cohorts, with missingness remaining below 2% and generally decreasing over time.

As the study progressed, the average time from session completion to PAC completion decreased to less than one day, indicating improvements in the timeliness of fidelity documentation over the course of the study. Collectively, these findings suggest that modifications to intervention delivery, Guide training, and fidelity monitoring procedures were associated with improved fidelity assessment reliability and documentation practices.

Participant protocol adherence was also monitored as part of study feasibility. Nine care partners were removed from the study by the principal investigator due to protocol deviations, the most common being failure to complete surveys within the required timeline.

Participant receipt and enactment data suggested that care partners engaged with and applied the core skills taught in Tele-STELLA. All participants completed at least one ABC plan, and most completed two. Importantly, on the Tele-STELLA experience survey, participants generally agreed with the statement, “I feel confident that I can use Tele-STELLA skills to manage upsetting behaviors” (mean = 4.10, range = 1–5).

Focus group findings provided additional insight into participant enactment of the ABC approach, with three themes emerging.

#### Understanding / Makes Sense

Understanding of the ABC approach varied and often developed over time. Some participants initially found the framework unintuitive or difficult to apply. Emotional overwhelm was often described as a barrier to initial understanding, with one participant noting they “had no concept… [they] could step back and think logically.” Over time, however, participants described the approach as “practical” and “easy to understand once I got past the emotional part.” These findings suggest that understanding of the intervention is mediated by emotional readiness.

#### Confidence Engaging / Using Skills

Participants described increased confidence in applying the ABC approach over time, particularly as they practiced and adapted the ABCs to their own situations. In many cases, the framework became internalized, allowing participants to use it more fluidly without relying on written materials. Participants described applying the approach reflexively, including to their own behavior (“I need to ABC it out for myself as well”). This shift suggests that confidence may develop as participants adapt the approach to their unique caregiving situations.

#### Ongoing Use

Participants’ descriptions of continued use of the ABC approach suggest that long-term feasibility depends on adaptation. While many did not continue using written materials or structured worksheets after the study, they reported continuing to apply the underlying logic of the intervention in more informal ways. This pattern indicates ongoing use may be supported when intervention components are internalized and integrated into everyday thinking rather than maintained as formal tasks.

To conclude, these findings indicate that the fidelity data captured both delivery of the intervention by Guides and participant receipt and enactment of core intervention skills over time. The formal architecture of the fidelity assessment allowed for nimble intervention when interventionists veered from the protocol, providing care partners with the appropriate “dose” of Tele-STELLA.

## 4 Discussion

Taken together, this paper addresses the efficacy, feasibility, acceptability, and fidelity of the Tele-STELLA intervention. Quantitative findings indicate that the Tele-STELLA intervention was effective in decreasing caregiver burden for those that participated in the main component, Nova. The improvement in burden was sustained across the booster component, Constellation.

The Nova component was feasible for the majority of those that completed it, and the experience survey indicates they were able to access the study. However, study participation became less feasible the longer care partners were in the study. The experience survey after Nova, completed by 76% of the full sample, revealed that most caregivers found the study acceptable. Qualitative data suggests that, while Tele-STELLA did not fit well for more experienced care partners, it was generally perceived as useful and beneficial.

The fidelity findings underscore the importance of building sound fidelity infrastructure into behavioral intervention trials from the outset, including standardized interventionist training, assessor calibration, documentation review, and feedback loops to monitor and prevent drift.

Our work diverges from typical reporting in the extant literature in that we provide a detailed report of the data related to feasibility and acceptability and we examined the relationship of dementia stage to attrition.^18–21^ We employed weekly surveys to collect high-frequency data on adverse events, an experience survey that garnered data pre-and post-intervention sessions, and focus groups to collect in-depth assessments of the participant experience. Our analytic approach allowed for identification of predictors of withdrawal based on common criteria. Taken together, the findings provide a full picture of attrition in this study.

Our study demonstrates effective management of the high attrition rate by adapting to the changing dynamics in the research milieu. Hagen et al. describe this as “kinesthetic learning,” in which recruitment efforts are altered based on emerging experience with the study.^5^ The inclusion and exclusion criteria of the Tele-STELLA study did not change, but we recognized early on that the attrition rate was higher than expected, threatening internal validity.^22^

The actual attrition rate of 44% provides a real-world example of the challenges scientists face when working with families in the later stages of dementia. The findings shed light on the relationship between dementia stage and study engagement. The Tele-STELLA sample was made up of care partners for those in the moderate to late stages of dementia, thus the risk of attrition increased over time due to increasing care demands in the context of progressive dementia. Because we did not include care recipients in the mild stages in the study, the number of care recipient deaths contributed to our high attrition rate.

Our experience suggests that, when enrolling care partners for those in the later stages of dementia, predicted attrition rates should range on the higher side. Further, the fidelity findings related to participant receipt and enactment suggest the potential value of assessing care partner emotional or learner readiness. Because some participants described emotional overwhelm as a barrier to initially understanding and applying the ABC approach, a brief readiness assessment may help identify care partners who need additional orientation, pacing, or support to fully engage with and enact core intervention skills. This may be particularly important in behavioral interventions for dementia care partners, where intervention uptake may depend not only on exposure to content, but also on the care partner’s emotional capacity to process and apply new strategies during stressful caregiving situations.

We recognize that the high number of deaths, health events, and care partner time demands drove our attrition rate up. However, behavioral symptoms occur across the disease trajectory and behavioral interventions, even in the late stages, can improve care partner well-being and decrease the number of health-related events.^23^ From an ethical standpoint, intervention testing needs to include care partners caring for those in the late stages, where burden is high. Excluding these care partners jeopardizes external validity—interventions that do not include families in the late stages cannot be generalized to the wider caregiving community.

## 5 Limitations and Strengths

Several limitations of this study are notable. First, the focus of this paper is on feasibility and acceptability. Limited efficacy data is provided for context, but a full report on efficacy is not described here.

Second, during the study planning period our original estimation of the attrition rate was too low (15%), necessitating an increase in sample size during implementation. We estimated initial rate based on our pilot work in which the sample size was small, and on published studies reporting attrition rates, which ranged 10-28%.^24,25^ However, this underestimation aligned with Aim 1, fostering a detailed examination of the high attrition rate.

Third, Tele-STELLA’s attrition rate might have been lower if we excluded care partners whose family members with dementia were in the very late stages of dementia. However, at enrollment, of those whose family members with dementia died, most reported their family members were in the moderate stages of dementia. Neither the care partners nor the study staff predicted the deaths and thus, the attrition rate would have only marginally improved if we had excluded care partners whose family members were in the late stages of dementia.

Several strengths of this study are apparent. First, we collected weekly data on adverse events, allowing us to link these events to feasibility and attrition. This detail is rarely seen in caregiver studies. Another strength of the study was the team’s early recognition that fidelity procedures required substantial refinement while the study was still underway. The revised fidelity procedures allowed the team to clarify protocol expectations, retrain interventionists and assessors, improve documentation processes, and monitor for drift across cohorts. The observed improvements in fidelity assessment reliability and documentation practices suggest that structured fidelity monitoring can be strengthened during active implementation and integrated into behavioral intervention delivery without undermining feasibility. However, this experience also highlights the importance of incorporating fidelity procedures from the outset in future trials.

Taken together, we recognize that the Tele-STELLA study had a high attrition rate (44%), nearly three times our original estimation and almost double that of other caregiver studies.^23^ We analyzed the qualitative and quantitative data to fully understand this phenomenon. Our results indicate that enrolling care partners whose family members are in the late stages of dementia risks high attrition rates due to care partner overwhelm and care recipient death. However, strategies described by Hagen et al. (2011)^5^ informed our current and future work. Further, we learned that our participants found that intervention helpful and meaningful, even when they couldn’t stay in the study. Our findings can inform others who embark on efforts to test interventions for families in the advanced stages of dementia.

## Data Availability

All data produced in the present study are available upon reasonable request to the authors.

https://www.ohsu.edu/alzheimers-disease-research-center/data-resources

## Acknowledgements

Tele-STELLA was funded by the National Institute on Aging (NIA), R01AG067546. Staff time was supported by NIA P30AG066518. The study was approved initially by Western IRB (#1303009). In 2022, IRB oversight was transferred to Oregon Health & Science University (IRB#22288). Tele-STELLA is registered with Clinicatrials.gov (#NCT04627662). The funders did not contribute to design, conduct, analysis and reporting of the trial, other than discussed above.

The authors are grateful for all the care partners who joined Tele-STELLA, whether or not they could stay in the full study. Your engagement was elemental to the success of the study-thank you.

## Conflict of Interest Statement

The authors declare that they have no known competing financial interests or personal relationships that could have appeared to influence the work reported in this paper.

## Consent Statement

All study participants, or their legally authorized representatives, provided informed verbal consent prior to study enrollment.

## Artificial Intelligence Statement

No generative AI tools were used in the creation of this manuscript. All research, writing, and analysis were performed independently by the authors.

## Notes

**Funding Information:** Tele-STELLA was funded by the National Institute on Aging (NIA) of the National Institutes of Health, Grant/Award Number: R01AG067546. Dr. Lindauer is funded by NIA R01AG067546. Staff time was supported by NIA P30AG066518, as are all co-authors. Dr. Cloyes is funded by NIA P30AG066518.

### Competing Interest Statement

The authors have declared no competing interest.

### Clinical Trial

NCT04627662

### Author Declarations

The study was approved initially by Western Institutional Review Board, a single Institutional Review Board, to oversee multiple sites (#1303009). In 2022, oversight was transferred to Oregon Health & Science University Institutional Review Board (l#22288). Tele-STELLA is registered with Clinicatrials.gov (#NCT04627662).

